# Unique combination of Recombinase Polymerase Amplification with label-free citrate-stabilised silver nanoparticle assay offers a highly sensitive, fast, accurate and visual molecular detection of *Klebsiella pneumoniae*

**DOI:** 10.1101/2024.01.17.24301463

**Authors:** Naresh Patnaik, Nidhi Orekonday, Ruchi Jain Dey

## Abstract

Our study addresses the growing concern posed by *Klebsiella pneumoniae*, a significant pathogen acknowledged by the World Health Organization (WHO). This bacterium is particularly alarming due to its association with antimicrobial resistance (AMR), impacting immunologically vulnerable populations, especially in hospital settings, and playing a crucial role in wound management. Moreover, this pathogen raises significant concerns in maternal and child health, being correlated with adverse outcomes like pre-term birth, low birth weight, and increased susceptibility to infections in new-borns, often resulting in morbidity and mortality.

A major obstacle to the effective and timely management of *K. pneumoniae* infections is the absence of rapid and cost-effective detection tools in resource-poor point-of-care (POC) settings. This study introduces an innovative combination of three POC-compatible methods: Insta DNA™ card-based sample collection and DNA extraction, Recombinase polymerase amplification (RPA)-based isothermal amplification, and a silver nanoparticle (AgNP) aggregation assay for visual detection. Together, these methods offer simple yet highly sensitive, specific, and rapid visual detection of as few as ∼1 bacterium of *K. pneumoniae* within ∼45 minutes. The synergy of these methods eliminates the need for sophisticated equipment, making it highly suitable for field and resource-poor POC applications.

## Introduction

The escalation of antimicrobial resistance (AMR) constitutes a significant global health threat, particularly within the notorious ESKAPE group of pathogens. This acronym encompasses six highly virulent and antibiotic-resistant bacterial pathogens: *Enterococcus species (spp.), Staphylococcus aureus, Klebsiella pneumoniae, Acinetobacter baumannii, Pseudomonas aeruginosa, and Enterobacter spp*. The ESKAPE group of pathogens has garnered its name owing to their characteristic ability to escape or circumvent the action of antimicrobial agents^1^. Within the ESKAPE group of pathogens, the World Health Organization (WHO) recognizes *K. pneumoniae* as a significant pathogen of concern, primarily due to its association with increasing AMR^1–3^. *K. pneumoniae* is recognized as a cause of nosocomial infection and can infect a variety of anatomical sites in the human body, causing severe conditions such as necrotizing pneumonia, urinary tract infections (UTIs), wound or surgical site infections, bloodstream infections, septicaemia, meningitis, and pyogenic liver abscesses, making it difficult to treat in healthcare settings^4–6^. Typically, immune-competent individuals do not succumb to *Klebsiella* infections. However, instances of *Klebsiella* infections are prevalent among patients undergoing treatment for other medical conditions such as COVID-19^7^. Those particularly vulnerable or immune-compromised including individual’s dependent on medical devices such as ventilators (respiratory machines) or intravenous catheters and those subjected to prolonged courses of specific antibiotics are often at heightened risk of contracting *Klebsiella* infections^8^.

*Klebsiella* infections are also important in the context of maternal and child health. In pregnant women, *Klebsiella* infections can contribute to complications such as chorioamnionitis, thereby increasing the likelihood of pre-term labour/birth^9^. Infections during pregnancy poses significant risk of vertical transmission to foetus or neonatal infections during or following the birth, leading to increased infant mortality^10, 11^. Hence, preventive measures and early detection of *Klebsiella* is paramount for timely intervention and effective management of these infections in maternal and child healthcare settings.

Early intervention facilitates prompt initiation of appropriate medical measures, reducing the risk of severe complications and preventing transmission. Conventional methods for detecting *K. pneumoniae*, such as culture-based and biochemical techniques, are time-consuming and provide limited information on abundance and strain diversity within samples, with uncertain sensitivity^12^. The recent introduction of whole metagenomics sequencing (WMS) in clinical diagnostics has significantly enhanced diagnostic reliability, particularly in intricate situations such as tissue biopsies and body fluids^12, 13^. WMS provides superior capabilities compared to traditional culture methods, offering comprehensive insights into antibiotic resistance profiles, virulence features, and evolutionary analysis of bacterial strains^12, 14^. Nevertheless, the exorbitant cost, time involvement and requirement of highly skilled individuals to perform and analyse the results currently restricts its routine application in diverse healthcare settings. In contrast, PCR-based assays, particularly real-time quantitative PCR^15^, have shown 100% sensitivity and specificity, with a limit of detection of 1-10 bacteria **(Table 1)**. However, the cost of detection is extremely high due to requirement of fluorescent probes and quantitative PCR (qPCR) equipment. Subsequent developments introduced isothermal amplification methods, including loop-mediated isothermal amplification (LAMP)^16, 17^ and recombinase polymerase amplification (RPA)^18–21^ and multiple cross displacement amplification (MCDA)^22^, for rapid *K. pneumoniae* detection without the requirement of expensive thermal cyclers or qPCR. LAMP alone or in combination with clustered regularly interspaced short palindromic repeats (CRISPR) and MCDA combined with lateral flow strip (LFS) has demonstrated comparable sensitivity (1-20 bacteria) compared to qPCR. These methods achieve detection within 60-90 minutes under isothermal conditions (∼60-65°C)^16, 17, 22^ **(Table 1).** In comparison, RPA provides further advantage of rapid (15-30 min) isothermal amplification at lower temperatures (∼37-42°C) permitting quicker detection^23^. RPA when combined with Lateral Flow Strip (LFS) system is capable of detecting as low as 100-1000 bacteria, highlighting scope for improving the sensitivity of detection post RPA-based DNA amplification^18, 19^. A recent study highlights utility of tailor-made plasmonic aptamer-gold nanoparticle (AuNP) assay for detection of Klebsiella without the need of amplification, however, the sensitivity of this methods is ∼3400 bacteria^24^. Despite the progress in molecular detection methods for *Klebsiella*, high cost associated with fluorescent primer probes, aptamers, additional reagents like CRISPR, LFS and AuNP poses accessibility challenges for low-income and resource-poor settings highlighting need for cost-effective visual detection tools. Another hurdle in POC settings is safe methods of sample collection, storage, transport and genomic DNA extraction for molecular detection. InstaDNA card consists of a special type of filter paper impregnated with a proprietary formula containing reagents that promote cell lysis and protein denaturation with subsequent release of nucleic acids that are entrapped within the matrix of the card and stabilized at room temperature, allowing long-term storage^25–27^. Originally applied to the perseveration of blood samples for forensics tests, collection of materials for genotypic analysis of microorganisms, plants, and molecular epidemiologic studies, the cards have also been used to store gynaecologic and non-gynaecologic cytology specimens for various molecular analyses^28^. Many advantages have been described for Insta DNA card, including low-cost, simple extraction protocols, easy transportation, minimal storage space and no special infrastructure being required^28^. However, there are no previous reports for application of InstaDNA cards for *K. pneumoniae* detection.

**Table 1.** List of visual detection methodologies developed previously for the molecular detection of *K. pneumoniae*.

| Sl. No. | Method of Amplification | Method of Detection | Sensitivity | Time complexity of detection | Reference |
| --- | --- | --- | --- | --- | --- |
| 1 | PCR | AGE | $1.2 \times 10^2$ bacteria | ~4-5 hours | 34 |
| 2 | Real-time PCR | Fluorescence | 1 bacterium | ~2 hours | 15 |
| 3 | Real-time PCR | Fluorescence resonance energy transfer (FRET) | 10 bacteria | ~2 hours | 35 |
| 4 | PCR | Gold nanoparticles | $16 \times 10^1$ bacteria (1pg gDNA) | ~2 hours | 24 |
| 5 | LAMP | AGE | 1 bacterium | ~25 min (LAMP) + 1hour Agarose gel | 16 |
| 6 | LAMP | CRISPR-based detection | $16 \times 10^1$ bacteria (1pg gDNA) | ~ 60 min | 17 |
| 7 | MCDA | Gold nanoparticle-LFS | 16 bacteria (100 fg gDNA) | ~ 30 min (estimated, not reported) | 22 |
| 8 | RPA | LFS | $10^3$ bacteria | ~15 min | 18 |
| 9 | RPA | LFS | $16 \times 10^3$ bacteria (0.1 ng/ul) | ~33 min | 19 |
| 10 | Real-time RPA | Fluorescence probes | 100-1000 bacteria | ~12 min | 20 |
| 11 | RPA | AGE | $10^2$ -$10^3$ bacteria | ~1 hour | 21 |

This study combines three unique POC friendly methods (a) InstaDNA card, for easy sample collection and DNA extraction (b) RPA-based thermal amplification in addition to PCR (c) application of silver nanoparticle (AgNP) aggregation assay developed in our laboratory^29^ for rapid, sensitive and visual molecular detection of *K. pneumoniae*. This unique combination is particularly suitable for *K. pneumoniae* detection in resource-poor settings without the need of costly thermal cycler and equipment like centrifuge or qPCRs. The integration of these methods is anticipated to address the clinical needs to mitigate the impact of *Klebsiella* infections.

## Results and Discussion

### InstaDNA card-based sample collection and DNA extraction followed by PCR based molecular identification of *K. pneumoniae*

We first optimized *K. pneumoniae* PCR, utilizing genomic DNA extracted using standard kit-based method and 2 sets of primers (**Table 2**), one described previously^30^ and another newly designed in this study. Both the primer sets target *16s rRNA* gene, however yield amplicons of two different sizes, ∼658 bp (primer set 1) and ∼250 bp (primer set 2), respectively. In addition, these primers were found to be highly specific without any cross-reactivity towards non-target organisms **(Figure 1 A and B)**. However, both the primer sets showed specificity towards *Enterobacter spp.* in addition to *K. pneumoniae* **(Figure 1 A and B)**, owing to the fact that both these organisms belong to the same family with a negligible genomic difference in their *16S rRNA* region. AGE-based detection post-PCR amplification showed a sensitivity of detection of ∼33 bacteria (equivalent to ∼200 fg of gDNA of *K. pneumoniae*) (**Figure 2 A and B**).

**Table 2.**
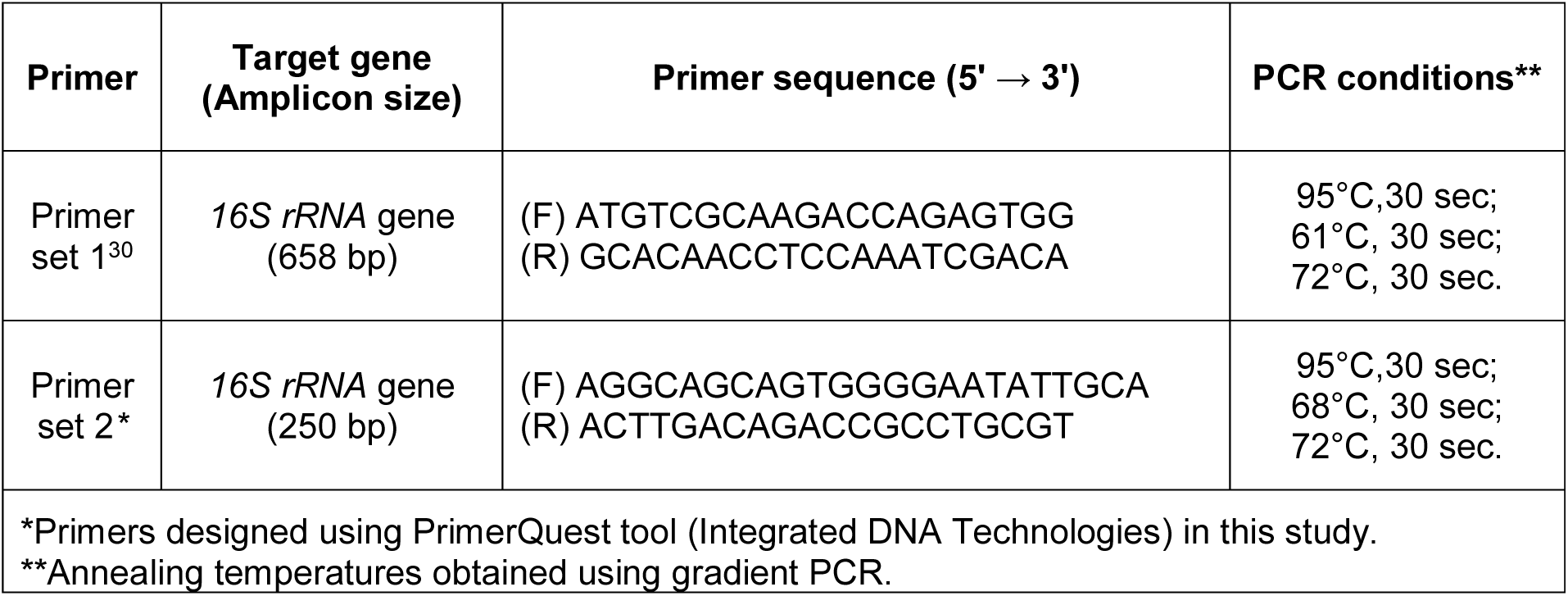
Primers for molecular detection of *Klebsiella pneumoniae*.

**Figure 1.**
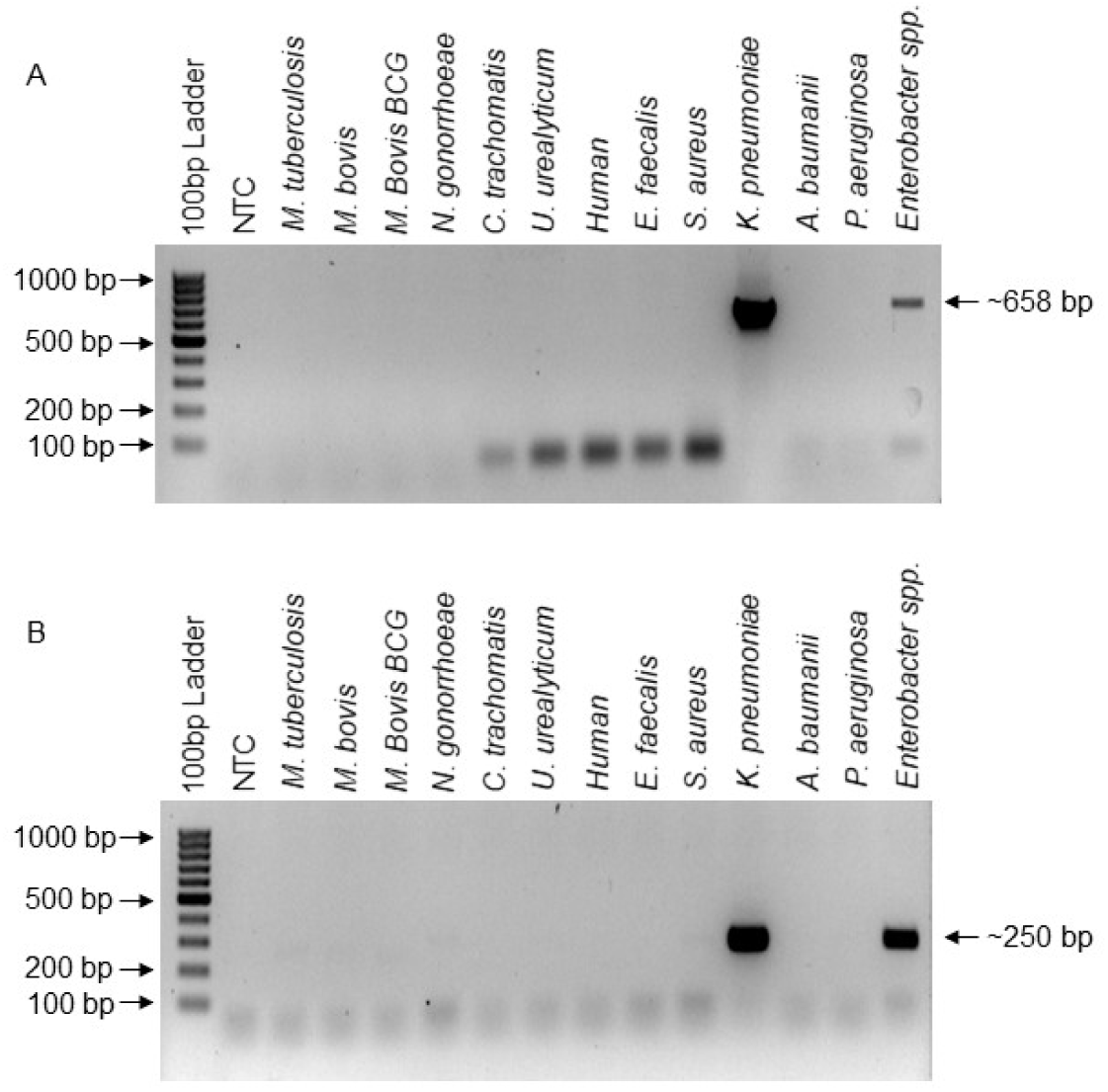
Primer specificity and cross-reactivity verification of primers utilized for detection of *K. pneumoniae* by PCR-AGE. Figures depict AGE-based verification of *K. pneumoniae* specific PCR carried out using two different primer sets designed to target *16S rRNA* gene, A. Primer set 1 produce an amplicon of ∼658 bp, and B. Primer set 2 produce an amplicon of ∼250 bp. The primer specificity was verified using genomic DNA derived from human and various human pathogens as negative controls such as, *M. tuberculosis*, *M. bovis*, *M. bovis* BCG, *N. gonorrhoeae*, *C. trachomatis*, *U. urealyticum*, and ESKAPE pathogens.

**Figure 2.**
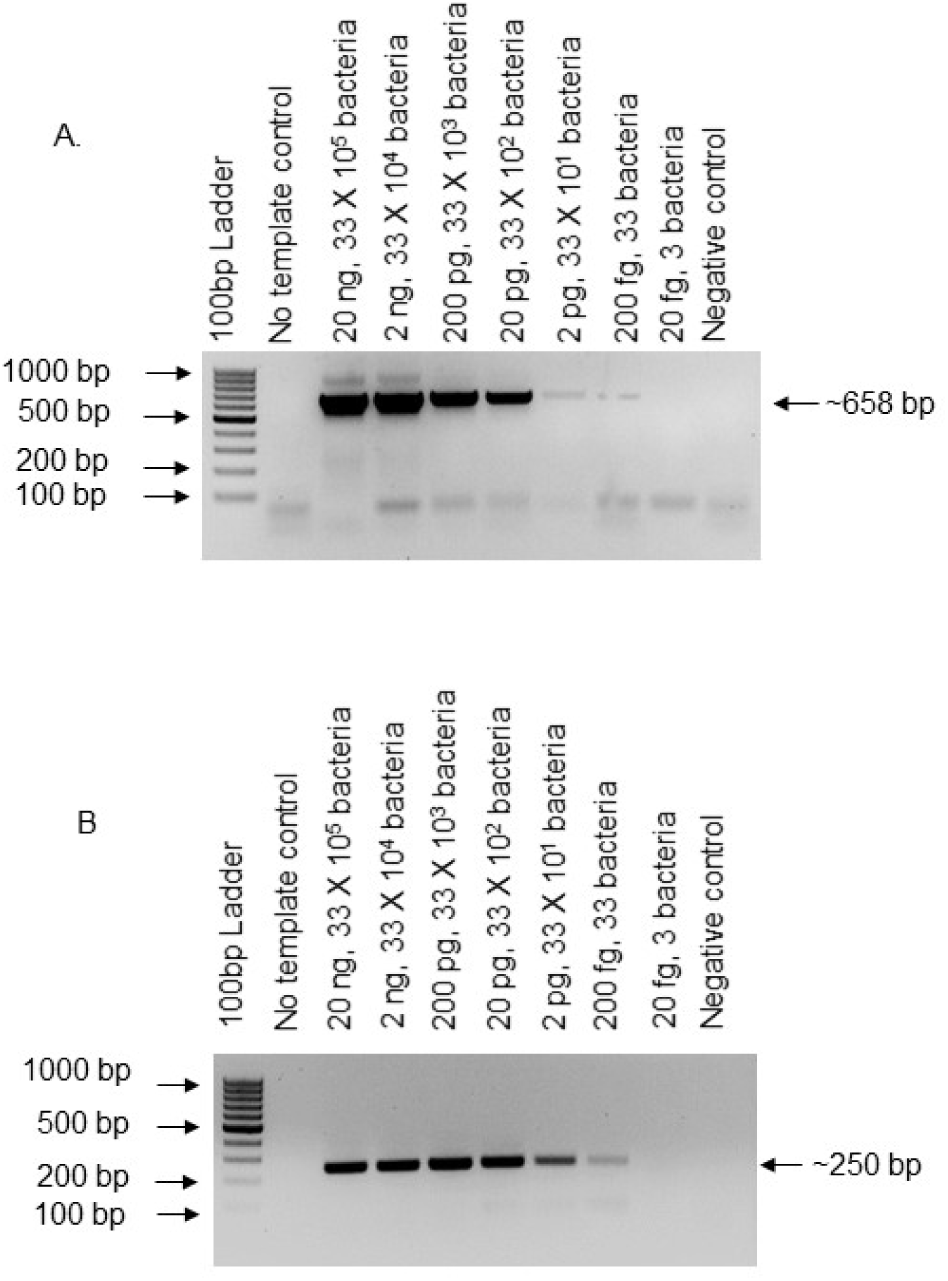
Sensitivity of *K. pneumoniae* PCR-AGE-based detection. Figure depicts a sensitivity of detection of ∼200 fg of gDNA of *K. pneumoniae* (∼33 bacteria) using PCR-AGE employing A. Primer set 1 (658 bp amplicon), and B. Primer set 2 (250 bp). The labels on top of the gel indicate amount of the genomic DNA used per PCR reaction & approximation of the number of bacteria. The negative controls include-No template control (NTC) and a mix of genomic DNAs obtained from human and human pathogens, such as, *M. tuberculosis*, *M. bovis*, *M. bovis* BCG, *N. gonorrhoeae*, *C. trachomatis*, *U. urealyticum*, and ESKAPE pathogens, except that of the test organism.

We next tested various genomic DNA isolation methods (boiling lysis, InstaDNA card and Whatman filter paper No. 1), suitable for POC using primer set 1 against 16S *rRNA* gene of *K. pneumoniae* **(Figure 3)**. The InstaDNA card and Whatman filter paper No. 1 methods show comparably identical efficiency in elution and amplification (Figure 3). Due to the advantages of InstaDNA cards pertaining to their ease of sample collection, storage and logistics, we selected InstaDNA card for subsequent analysis and determined their sensitivity of detection for *K. pneumoniae*. The synergy of InstaDNA card followed by PCR-AGE shows a sensitivity of ∼330 bacterial cells of *K. pneumoniae* **(Figure 4)**.

**Figure 3.**
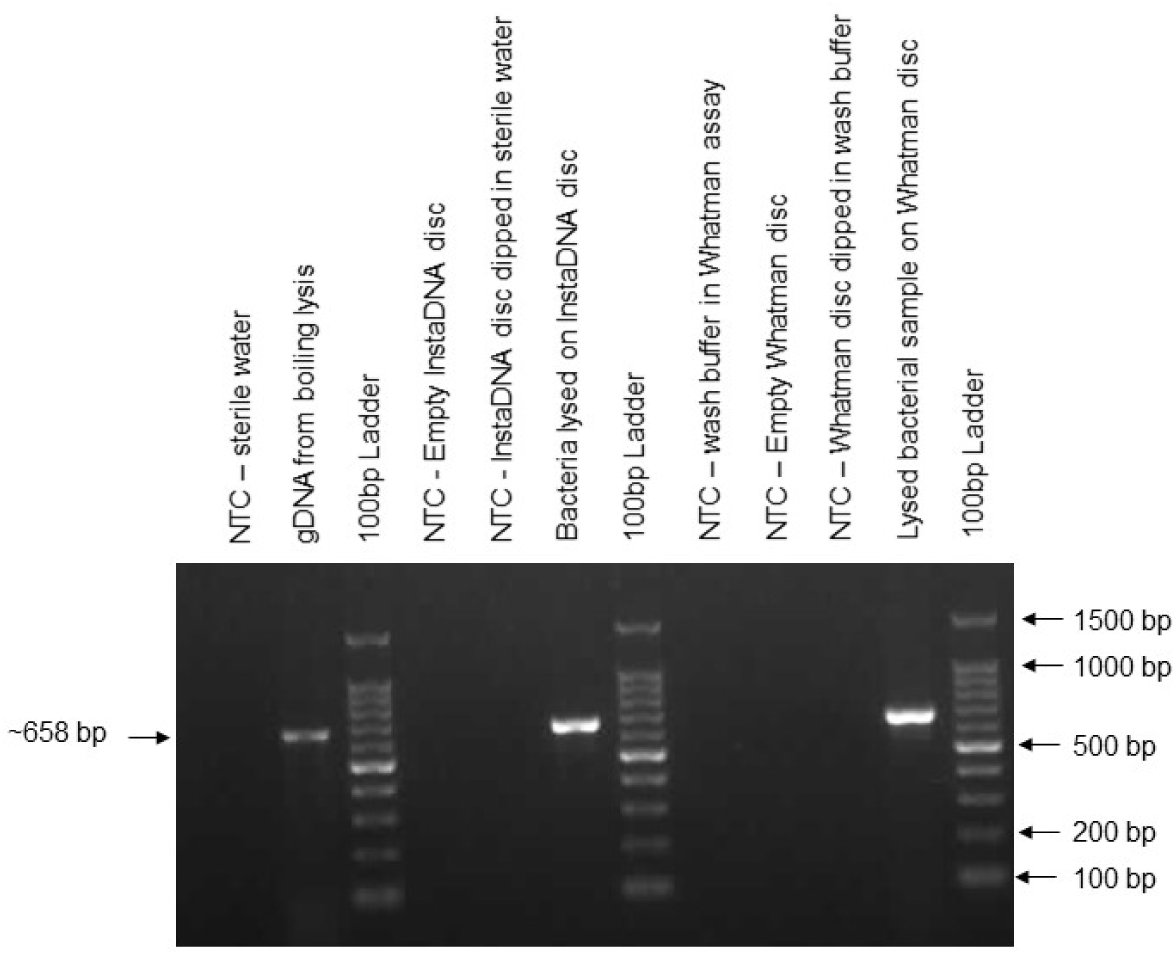
Comparative analysis of various POC gDNA isolation techniques using PCR-AGE-based detection. Figure depicts the AGE-based detection of PCR performed on gDNA isolated using different POC methodologies, namely, boiling lysis, InstaDNA card-based isolation, and Whatman Paper No 1 assay. The PCR was verified using primer set 1 (∼658 bp) specific to *16S rRNA* gene of *K. pneumoniae*. InstaDNA card-based DNA isolation and Whatman Paper No 1 assay displayed comparable efficiency of DNA isolation. The labels on top of the gel indicate amount of the genomic DNA used per PCR reaction and the negative controls used for each technique.

**Figure 4.**
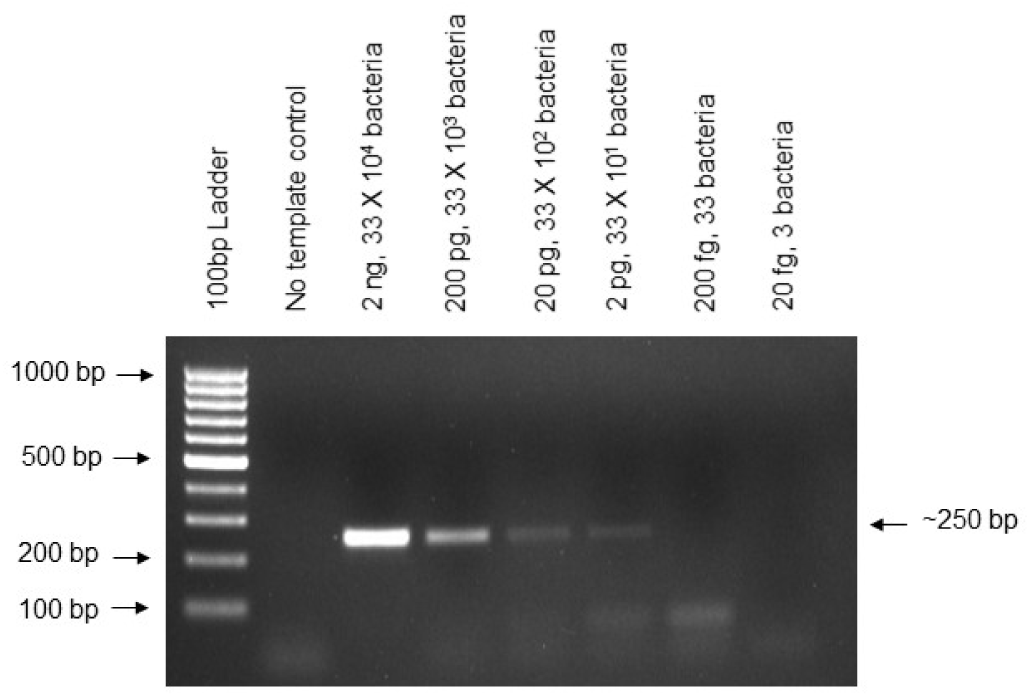
Sensitivity of *K. pneumoniae* detection using InstaDNA card-based DNA isolation followed by PCR-AGE for detection. Figure depicts the AGE-based detection of PCR performed on various dilutions of bacterial sample blotted onto InstaDNA card disc, without elution step, and using primer set 2 (∼250 bp) specific to *16S rRNA* gene of *K. pneumoniae*. The assay shows a sensitivity of detection of ∼2 pg of gDNA of *K. pneumoniae* (∼330 bacteria). The labels on top of the gel indicate amount of the genomic DNA used per PCR reaction & approximation of the number of bacteria. The negative controls included a No template control (NTC).

### Bridging flocculation assay and AgNP assay for molecular detection of *K. pneumoniae* post-PCR amplification

We next performed paramagnetic bead-based bridging flocculation assay following PCR and determined the sensitivity of visual detection and compared with PCR-AGE. PCR performed using both the primer sets (**mentioned in Table 2**) showed a comparable detection sensitivity of ∼33 bacteria **(Figure 5 A and B)**, which is similar to PCR-AGE (**Figure 2 A and B**).

**Figure 5.**
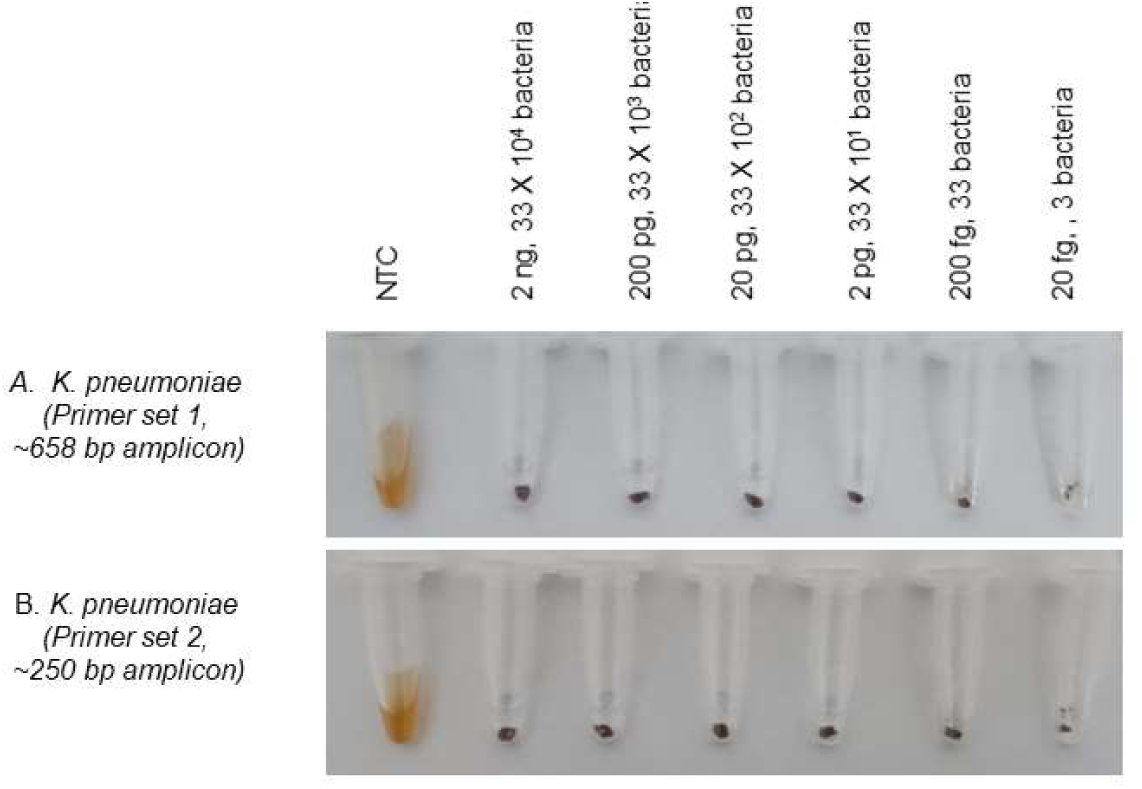
Sensitivity of *K. pneumoniae* detection by PCR and paramagnetic bead-based bridging flocculation assay. Paramagnetic bead-based bridging flocculation assay was performed on PCR products obtained using 16S rRNA gene-specific primer set 1 and 2 yielding amplicon size of A. 658 bp and B. 250 bp, respectively. The assay displays a similar detection sensitivity of 200 fg (∼33 bacteria) using both the primer sets. Presence of PCR amplified DNA results in a visual floc (bead aggregate), however, absence or very low amount of amplified DNA result in dispersed state of magnetic beads (brown solution). The sensitivity of PCR-AGE (Figure 2) and PCR-Bridging flocculation assays were found to be comparable.

For further enhancing the sensitivity of visual detection, we performed AgNP assay on DNA obtained from PCR. The AgNP-based assay showed a 10-fold higher sensitivity of detection (∼3 bacteria, equivalent to ∼20 fg of gDNA of *K. pneumoniae*) compared to PCR-AGE and bridging flocculation (**Figure 6 A and B**). Hence, post-PCR amplification, silver nanoparticle assay, exhibits a higher sensitivity of detection as compared to flocculation assay and traditional AGE-based detection.

**Figure 6.**
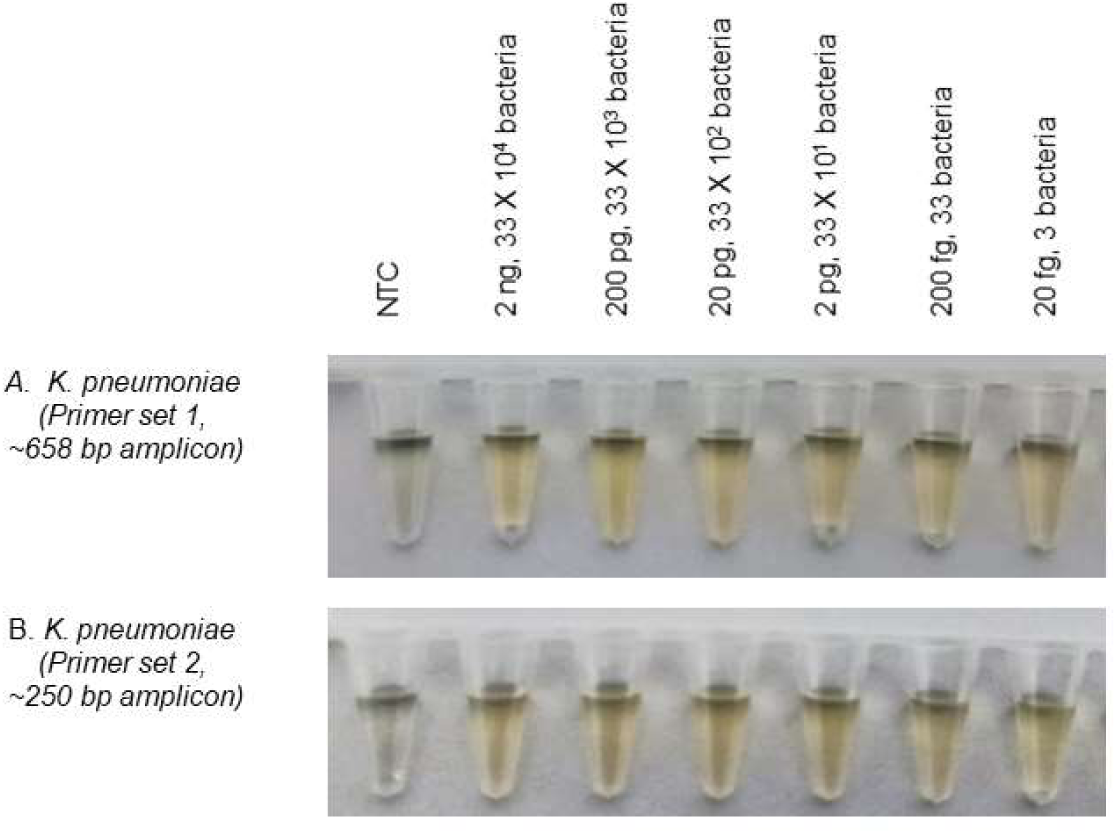
Sensitivity of detection of *K. pneumoniae* by PCR and AgNP assay. AgNP aggregation-based visual detection assay performed on PCR products obtained using 16S rRNA gene specific primer set 1 and 2 producing amplicon of, A. 658 bp and B. 250 bp, respectively. Both the primer sets display detection sensitivity of 20 fg of *K. pneumoniae* genomic DNA (∼3 bacteria). Presence of amplified DNA causes AgNPs to remain in dispersed state indicated by yellow color solution, however, absence or very low amount of amplified DNA results in aggregation of AgNPs in presence of the aggregating agent (NaCl), indicated by grey color solution.

### RPA-based Isothermal amplification of *K. pneumoniae 16S rRNA* region followed by in-tube detection using AgNP

We next assessed if isothermal amplification techniques such as RPA can be used in combination with AgNP assay for rapid molecular detection of *K. pneumoniae*. RPA performs amplification in ambient conditions (25 - 42°C) and hence well suited for resource poor point-of-care (POC) settings, where scope of costly equipment is rare. We performed the RPA amplification of the target regions of *K. pneumoniae* at 37°C. Out of the two primer sets designed for the molecular detection of *K. pneumoniae*, only primer set 2 was found suitable for consistent amplification using RPA. RPA-AGE showed a dramatic 33-fold improvement in sensitivity of detection (∼1 bacterium) **(Figure 7 A)**, compared to PCR-AGE (∼33 bacteria) (**Figure 2 B**). RPA-AgNP assay **(Figure 7 B)** also exhibited marked improvement in sensitivity of detection to as low as 1 bacterium comparable to RPA-AGE **(Figure 7 A)** and was also comparable to PCR-AgNP assay **(Figure 6 B)**. We also tested paramagnetic bead-based bridging flocculation-based visual detection in combination with RPA. While the beads offer an easy method to clean up RPA amplified products, however, gives inconsistent results when used for flocculation based visual detection, hence not utilized for detection purpose (data not shown).

**Figure 7.**
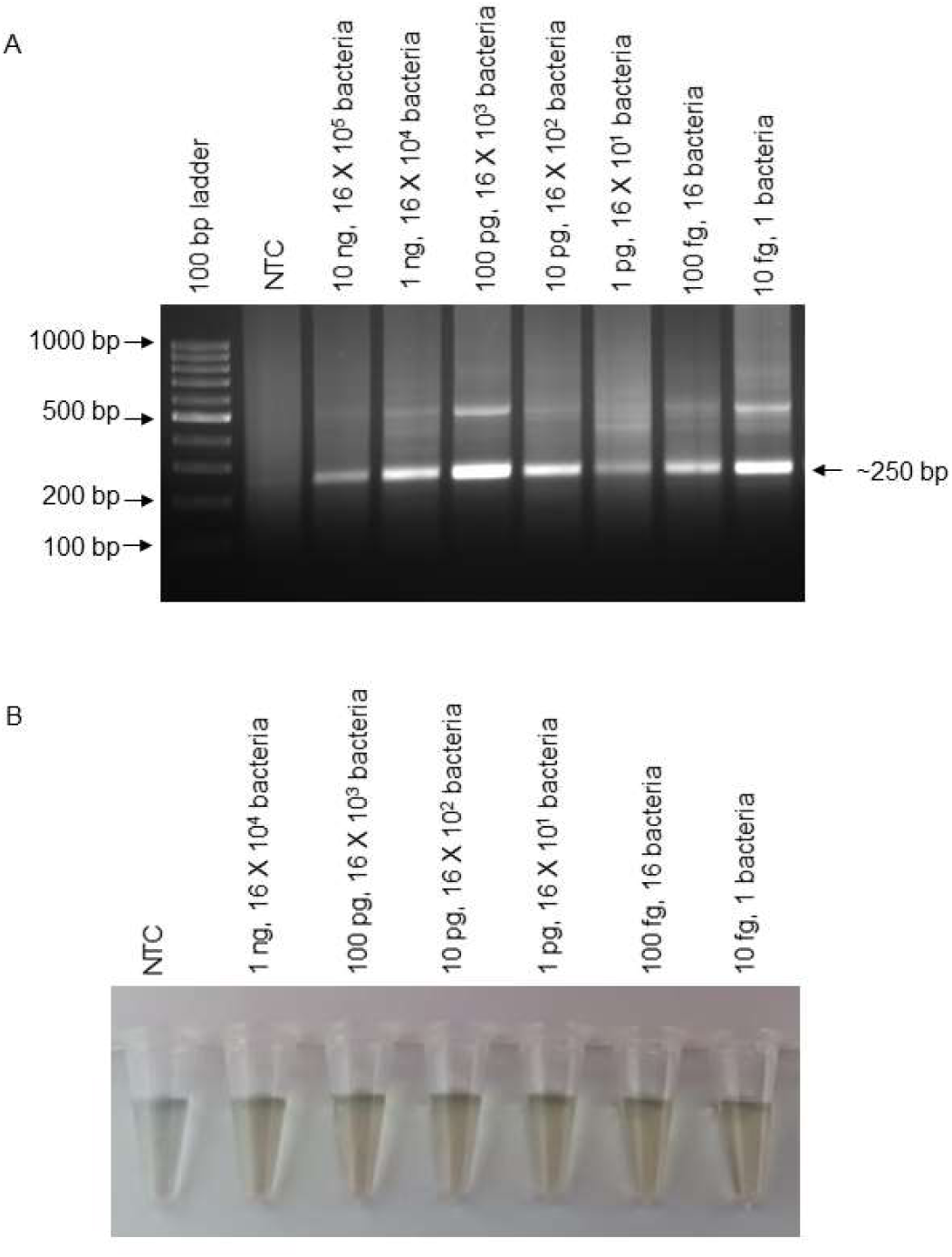
Sensitivity of detection of *K. pneumoniae* by RPA-AGE and RPA-AgNP assay. Figure depicts, A. AGE-based and B. Ag-NP assay-based detection of DNA amplified using RPA employing primer set 2 (amplicon size, ∼250 bp). The labels on top of the gel indicate amount of genomic DNA used per RPA reaction and the corresponding number of bacteria. Isothermal RPA-AGE and RPA-AgNP assays show comparable detection sensitivity of ∼10 fg of *K. pneumoniae* genomic DNA, equivalent to ∼1 bacterium.

## Conclusion

This study offers a highly sensitive, rapid method for molecular detection of an important human pathogen *K. pneumoniae* employing a unique combination of Insta-DNA card-based DNA extraction, isothermal DNA amplification using RPA and AgNP assay for visual detection. The RPA technique for isothermal DNA amplification, distinguishes itself from PCR by the virtue of its speed of operation (15-20 min) at a constant temperature range (∼25 - 42°C) compared to other isothermal techniques like LAMP and MCDA requiring more time (∼30 – 60 min) and higher temperature (60 - 65 °C). AgNP-based assay reported by us previously for differential molecular detection of *Mycobacterium tuberculosis* and *M. bovis*^29^, offers unique advantage of visual detection in a highly sensitive and cost-effective manner. The synergy of RPA and the AgNP assay eliminates the need for expensive thermal cycling equipment, centrifuges, electrophoresis equipment, gel documentation facilities, and real-time qPCR machines, making RPA particularly suitable for field and POC applications. Additionally, the primer sets designed in this study allow the application of the same primers for both PCR and RPA-based assays. However, considerations during primer design include a higher GC content (≥ 50%), longer primer length (20-35 bp), and maintaining the amplicon size between 200-500 bp. These primer sets are non-labelled, hence further keeping the cost of detection under check. This study is also the first report for application of paramagnetic bead-based bridging flocculation assay for highly sensitive visual detection of *K. pneumoniae* subsequent to PCR based amplification. InstaDNA card further offer a safe and simple method for sample collection, storage and transport and rapid DNA extraction without need of any sophisticated equipment or DNA extraction kits in POC settings in a highly cost-effective manner (< 1 $). Moreover, the DNA isolated using this method, is compatible with down-stream amplification methods, though slightly less sensitive (∼10 fold).

For the application of RPA based molecular detection with AgNP assay, it is important to include a clean-up process to eliminate the dense reagents like PEG and RPA enzymes which interferes with AGE and AgNP assay. Our study utilizes a highly efficient, yet cost-effective and equipment-free method based on paramagnetic beads for PCR and RPA derived amplicon clean-up process. The total cost of molecular detection utilizing a combination of three POC methods is <5 $ for InstaDNA-RPA-AgNP in comparison to <3$ for InstaDNA-PCR-AgNP (including the cost of paramagnetic beads used for clean-up)^31^. The only limitation of the RPA components at present is the higher cost compared to the PCR reagents, however, considering the suitability of RPA to POC settings, efforts to mass produce these reagents at lower cost may help in increasing the application of this useful tool in clinical settings and make it more robust and affordable compared to PCR/qPCR^31^. Incorporation of such isothermal reactions can significantly enhance the efficiency of diagnostic procedures, facilitating a streamlined and expedited process for the accurate and timely recognition of important pathogens in POC healthcare settings.

## Materials and methods

### Ethics Statement

The study was performed as per the guidelines and approval of the Institutional Biosafety Committee (BITS/IBSC/2019-1/7 and BITS/IBSC/2019-1/8) utilizing appropriate biosafety level 2 measures.

### Reagents and bacterial strains

All the reagents employed in this study were sourced from Himedia Laboratories Pvt. Ltd., unless mentioned otherwise. The oligonucleotide primers utilized, as outlined in **Table 1**, were obtained from Sigma Aldrich. MagGenome supplied the paramagnetic beads, while Sigma Aldrich provided the AgNP (20 nm size, stabilized in citrate buffer, 20 mg/ml). Taq polymerase and its associated reagents were procured from Takara Clontech. Twist Amp Liquid Basic RPA Kit was obtained from TwistDX^TM^.

Cultures of *Enterococcus faecalis* (ATCC 29212), *S. aureus* (MTCC 96), *K. pneumoniae* (MTCC 109), *A. baumanii* (MTCC 12889), *P. aeruginosa* (MTCC 424), *Enterobacter spp*. (MTCC 7104) and *Neisseria gonorrhoeae* (ATCC 19424) were acquired from the American type culture collection (ATCC, distributed by Himedia Laboratories Pvt. Ltd.) or Microbial Type Culture Collection and Gene Bank (MTCC), maintained by Institute of Microbial Technology (IMTECH), Chandigarh, India. The WHO BCG Danish 1331 vaccine sub-strain was obtained from the National Institute for Biological Standards and Control (NIBSC), UK.

### Sample Collection and DNA extraction

Genomic DNA (gDNA) was extracted from pure bacterial cultures using a standard kit-based method, specifically the DNEasy Blood and Tissue Kit (Qiagen), with minor adjustments to the manufacturer’s protocol. For cost-effective and rapid gDNA isolation, we employed various protocols, including Boiling Lysis^32^, Whatman filter paper No. 1^33^ and InstaDNA card method^25–27^. The resulting gDNA was quantified using NanoDrop Microvolume Spectrophotometers (Thermo Fisher Scientific) and utilized for amplification reactions with species-specific primers.

In the Boiling Lysis method, bacterial cell-pellets were resuspended in an in-house lysis buffer (1X TE, 0.05% Triton-X 100), boiled at 95°C for 30 minutes, and centrifuged at 12,000g for 10 minutes to obtain the genomic DNA-containing supernatant. For the Whatman paper method, microbial cells were washed with 1X PBS, resuspended in an in-house extraction buffer (2% w/v CTAB, 1.42 M NaCl, 20 mM EDTA, 100 mM Tris-HCl [pH 8.0], 1% w/v polyvinylpyrrolidone [PVP 40]), and lysed. A 3 mm Whatman No.1 disc was incubated in the cell lysate, washed, and directly transferred to the PCR reaction mix for amplification. Alternatively, DNA was eluted by dipping the disc in elution buffer (1X TE), heated at 95°C for 2 minutes, and quantified as described above. In the InstaDNA card method, a known bacterial culture of colony forming units (CFU), e.g., 33×10^4/2 µL, was blotted onto a 3 mm InstaDNA card disc, allowed to dry, washed, and transferred to the PCR reaction mix for amplification. Alternatively, the disc was transferred to elution buffer for DNA elution by heating at 95°C for 2 minutes. Similarly, 10-fold serially diluted bacterial samples were blotted onto separate discs and processed accordingly.

### PCR and Agarose gel electrophoresis (PCR-AGE)-based detection

Specific oligonucleotide primers targeting *K. pneumoniae* were custom-designed and are detailed in **Table 2**. Two sets of primers (set 1 and set 2) were designed targeting *16s rRNA* gene of *K. pneumoniae* to achieve an amplicon size of ∼658 base pairs and ∼250 base pairs, respectively. Primer specificity was confirmed *in silico* through NCBI-BLAST analysis. PCR amplification was carried out in a Veriti Thermal Cycler from Applied Biosystems, using Taq Polymerase sourced from Takara Clontech, following the manufacturer’s instructions with minor adjustments. Each 50 μl PCR reaction was prepared with a 1 μM primer mix in the reaction mixture, and thermal cycling included an initial denaturation step at 95°C for 10 minutes, followed by 35 cycles of PCR conditions outlined in **Table 2**. The process concluded with a final extension at 72°C for 10 minutes.

In order to validate the primer specificity for *K. pneumoniae*, genomic DNA samples from diverse organisms which could possibly cause respiratory infections or pneumoniae, were utilized as negative control, such as *Mycobacterium tuberculosis* (*M. tb*), *Mycobacterium bovis* (*M. bovis*), *M. bovis* BCG, *N. gonorrhoeae*, *Chlamydia trachomatis*, *Ureaplasma urealyticum*, *E. faecalis*, *S. aureus*, *A. baumanii*, *P. aeruginosa*, *Enterobacter spp* and human DNA. The verification of PCR products was performed through AGE and DNA was visualized by ethidium bromide dye-based staining. For sensitivity determination or limit of detection analysis of PCR-AGE, serial ten-fold dilutions of genomic DNA templates of *K. pneumoniae* ranging from 10 nanograms, ng (1.6×10^6^ bacteria) to 10 femtograms, fg (1 bacterium) were employed in PCR reactions.

### RPA-AGE-based molecular detection of *K. pneumoniae*

Isothermal nucleic acid amplification was performed for the pathogens using Twist Amp Liquid Basic RPA Kit (TwistDX, TALQBAS01kit) as recommended by the manufacturer with minor modifications. Briefly, all the reagents provided in the kit are mixed with defined quantity of DNA, reaction is initiated by addition of magnesium acetate, followed by incubation at 37°C for 15-20 minutes. The RPA products are immediately cleaned up using XpressPure PCR Cleanup kit (MagGenome, MG20Pcr-50) or NucleoSpin gel and PCR clean-up kit (Macherey-Nagel, 740609.250) to eliminate enzymes, highly dense PEG and buffer components which are known to interfere with the agarose gel electrophoresis. Both the kits allow equivalent performance in terms of quality of the isolated DNA, however, MagGenome kit allows faster (20 minutes) purification using simple magnetic stand-based purification unlike other PCR clean up kits needing centrifuge and ∼60 min purification time. Henceforth, we utilized instrument-free paramagnetic bead-based purification for our study. Similar to PCR-AGE assay, we determined the sensitivity or limit of detection of the RPA-AGE using serial ten-fold dilutions of genomic DNA templates (10 ng - 10 fg, equivalent to 1.6×10^6^ bacteria – 1 bacterium of *K. pneumoniae*). DNA was visualized by ethidium bromide dye-based staining.

### Visual detection of amplified products using bridging flocculation assay and citrate-stabilized AgNPs

Bridging flocculation assay and AgNP-based visual detection assays were performed using PCR amplified products as described by us previously^29^. Briefly, paramagnetic beads work with a principle that they adsorb the amplified DNA molecules and flocculate upon addition of flocculation buffer (sodium acetate and tween 20 at an acidic pH) due to the formation of DNA bridges connecting the paramagnetic beads. Hence, presence of DNA amplified product is determined by floc formation. Absence of DNA or lesser amount of the amplified product doesn’t form visible floc as DNA bridges are not formed, as indicated by brown dispersion^29^. Similarly, AgNP assay was carried out as described by us previously^29^, wherein presence of DNA prevents the aggregation of AgNPs upon addition of aggregating agent, sodium chloride (NaCl) causing sliver dispersion to maintain yellow colour. However, in absence of DNA, addition of NaCl leads to AgNP aggregation causing grey coloration of the dispersion.

While PCR products were detected using AGE, bridging flocculation and AgNP assay, RPA-amplified products were exclusively detected visually using AgNP assay given the higher sensitivity of AgNP-based visual detection (∼3 bacteria) of PCR amplified products compared to bridging flocculation (∼33 bacteria). Briefly, 10 μl of RPA reaction products were made up to a volume of 50 μl using sterile water. Amplified products were then cleaned-up using MagGenome PCR clean up kit as per the manufacturer’s instructions. Finally, DNA was eluted in 20 μl of elution buffer (10 mM Tris, 1mM EDTA, pH 8.0) followed by incubation with 50 μl of the citrate-stabilized AgNP dispersion at room temperature for ∼1 minute. Following this, 5 μl of aggregating reagent (5M NaCl) was added to the assay tubes. The onset of AgNP aggregation is promptly revealed (in < 1 min) by a grey colour dispersion in tubes lacking amplicons (negative controls with DNA from non-target organisms or no-template controls, NTC). Conversely, in tubes with amplified DNA, the addition of NaCl prevents AgNP aggregation, resulting in the retention of a yellow colour. The entire process of purification and visual detection is completed within a time frame of 20-25 minutes. The sensitivity of detection was assessed by executing AgNP aggregation-based assay using amplicons derived from RPA conducted with varying amounts of genomic DNA sourced from *K. pneumoniae*.

## Ancillary Information

### Author Contribution

NP, NO and RJD have conceived and designed the project. NP and NO have equal contribution to the work. NP and NO carried out the studies and designed figures and analysed the results. RJD received the funding. NP and RJD have written and edited the manuscript. All the authors reviewed and edited the manuscript.

### Funding

RJD is thankful to Birla Institute of Science and Technology (BITS) Pilani, Hyderabad campus, India for their funding support through intramural funding under Research Initiation Grant (RIG) and Centre for Human Disease Research (CHDR). RJD is thankful to Defence Research and Development Organization (DRDO), India for supporting through research grant (LSRB/81/48222/LSRB-367/BTB/2020). RJD is thankful to the overall infrastructure support by Department of Biological Sciences, BITS Pilani Hyderabad. NP is thankful to Indian Council of Medical Research (ICMR), Govt. of India for providing Senior Research Fellowship (RBMH/FW/2019/13). NO is thankful to LSRB, DRDO and BITS Pilani Hyderabad for fellowship.

## Acknowledgements

We acknowledge Dr. Raghunand R Tirumalai, Centre for Cellular and Molecular Biology (CCMB), Hyderabad, India for providing genomic DNA for *M. tuberculosis H37Rv.* Dr. Bappaditya Dey, National Institute of Animal Biotechnology (NIAB), Hyderabad, India is acknowledged for *M. bovis* genomic DNA. We are grateful to Dr. Benu Dhawan, Department of Microbiology, All India Institute of Medical Sciences (AIIMS, New Delhi, India) for giving genomic DNA from *Ureaplasma urealyticum*. We thank Dr. Karthika Rajeeve, Rajiv Gandhi Centre for Biotechnology, Trivandrum, India for providing *Chlamydia trachomatis* (L2 Serovar) genomic DNA.

## Data availability statement

All data that supports the findings of this study are available from corresponding author upon reasonable request.

## Abbreviations

*M. tb*: *Mycobacterium tuberculosis*
*M. bovis*: *Mycobacterium bovis*
POC: Point-of-care
ml: Millilitre
µl: microlitre
PCR: Polymerase Chain Reaction
DNA: Deoxyribonucleic acid
qPCR: Quantitative PCR
NTC: no-template-control
NaAc: Sodium Acetate
AgNP: Silver Nanoparticles
dsDNA: Double stranded DNA
Tris: Tris(hydroxymethyl)aminomethane
EDTA: Ethylene diamine tetra-acetic acid
NaCl: Sodium Chloride
LAMP: Loop-mediated Isothermal Amplification
RPA: Recombinase Polymerase Amplification
MCDA: Multiple cross displacement amplification
BCG: Bacille Calmette-Geurin
WHO: World Health Organisation.

## For Table of Contents Use Only

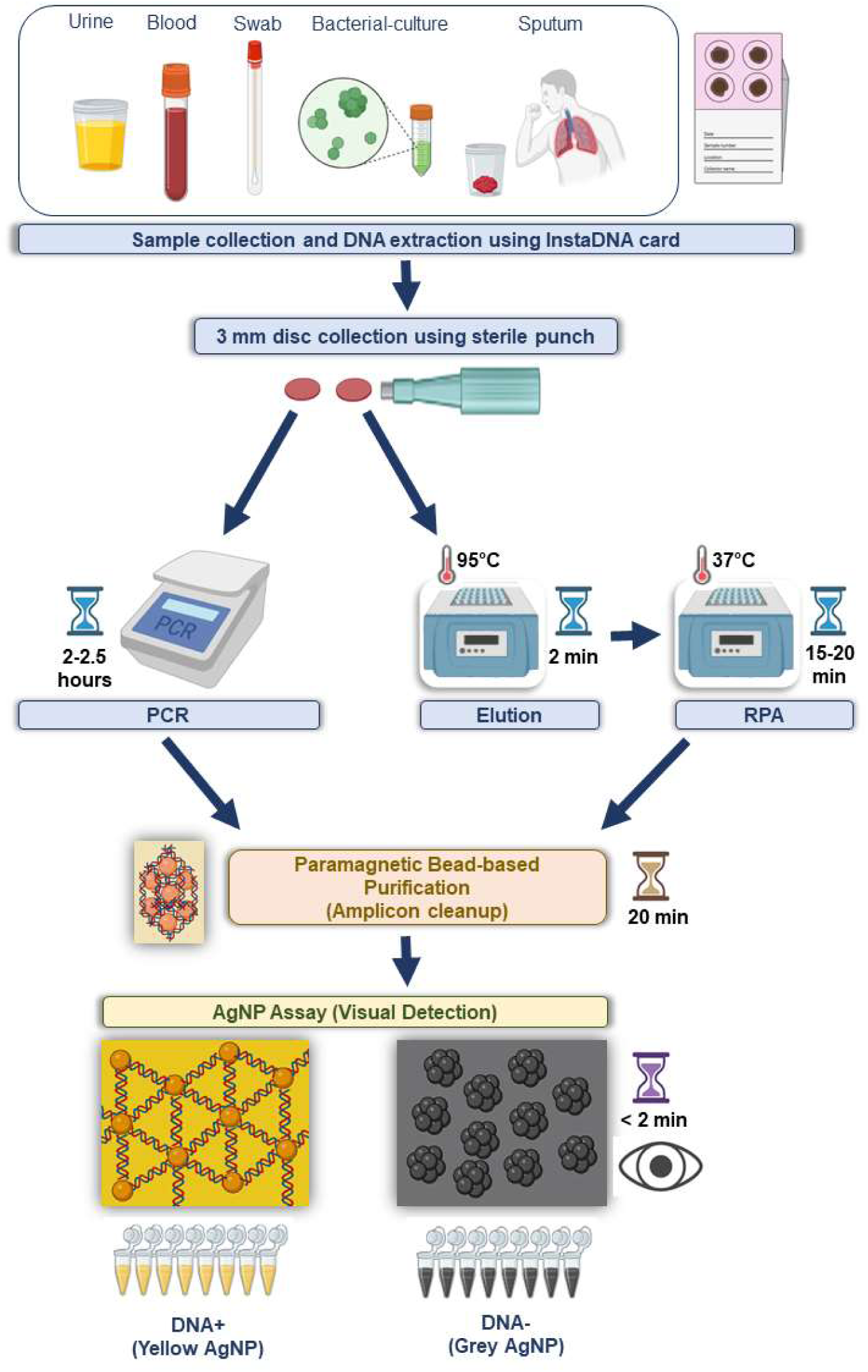

